# Pulmonary *FABP4* is an inverse biomarker of pneumonia in critically ill children and adults

**DOI:** 10.1101/2024.08.19.24312242

**Authors:** Emily C. Lydon, Hoang Van Phan, Eran Mick, Natasha Spottiswoode, Carolyn S. Calfee, Peter M. Mourani, Charles R. Langelier

**Author notes:** equal contributions (Emily C. Lydon, Hoang Van Phan). **Corresponding Author:** Emily C. Lydon, 513 Parnassus Avenue, Room S380, San Francisco, CA 94143. **Contributions:** CSC, PMM, CRL secured funding and designed and enrolled the clinical cohorts. ECL, CRL, PMM, CSC, and NS performed clinical adjudications for the cohorts. CRL performed RNA extractions and sequencing. EM, HVP, CRL, NS, and ECL conceived this specific project idea. HVP and EM performed data analysis and generated the figure. ECL, HVP, and CRL wrote the initial draft of the manuscript. All authors reviewed and approved the final manuscript. **Funding:** 5R01HL155418 (CRL, PMM); Chan Zuckerberg Biohub San Francisco (CRL); R35HL140026 (CSC); R01HL124103 (PMM).

## Abstract

Lower respiratory tract infection (LRTI) is a leading cause of mortality, yet accurate diagnosis remains challenging. We studied two prospective observational cohorts of critically ill patients with acute respiratory failure (pediatric, N=261; adult N=202) and analyzed RNA sequencing data from tracheal aspirate collected following intubation. Patients with LRTI or non-infectious acute respiratory failure were adjudicated by a multi-physician team. We found that *FABP4* expression was significantly downregulated in children with confirmed LRTI compared to those with non-infectious respiratory failure, and when incorporated into a diagnostic classifier, achieved an area under the receiver operating characteristic curve (AUC) of 0.90±0.07. When tested in the adult cohort, and AUC of 0.85±0.12 was attained. These findings suggest that pulmonary *FABP4* could serve as a valuable biomarker for the early diagnosis of LRTI in critically ill patients across the age spectrum, offering a potential tool for improving clinical outcomes and reducing unnecessary antibiotic use.

## Manuscript

Lower respiratory tract infection (LRTI) is a leading cause of death worldwide, yet the diagnosis is often challenging since non-infectious respiratory processes can present similarly. Furthermore, existing culture-based microbiologic testing is time-consuming, can return falsely negative with preceding antibiotics, and may identify microbes unrelated to a pulmonary infection. Consequently, clinicians often turn to rapid host response-based peripheral blood biomarkers, such as procalcitonin and C-reactive protein (CRP), for gauging LRTI suspicion and deciding to initiate empiric antibiotics. Procalcitonin has some utility in distinguishing bacterial and viral etiologies of LRTI, but diagnostic performance is limited in critically ill patients, and CRP performs worse.(1,2) Presently, no clinically available host biomarkers accurately distinguish all-cause LRTI from non-infectious respiratory illness.

To address the need for better LRTI diagnostics, we previously utilized respiratory metatranscriptomics to build multi-gene host classifiers that accurately differentiate LRTI from non-infectious acute respiratory syndromes in critically ill adults and children. These classifiers achieved remarkable accuracy, both when used on their own and when integrated with microbial detection.(3,4) While promising, the practical complexities of RNA sequencing precludes immediate implementation in many clinical settings. We therefore sought to identify whether a single gene could accurately diagnose LRTI in patients with acute respiratory failure.

We first evaluated a recently described(4) prospective multicenter cohort of 261 critically ill children with acute respiratory failure (Table 1). Transcriptional profiling was performed on tracheal aspirate (TA) samples collected <24 hours after intubation using previously described methods.(4) Retrospective clinical adjudication classified subjects into four LRTI groups: Definite (clinical LRTI diagnosis with compatible microbiology, n=117), Suspected (clinical LRTI diagnosis with negative microbiology, n=57), Indeterminate (clinical uncertainty regarding LRTI, n=37), and No Evidence (clear non-infectious etiology with negative microbiology, n=50). Most Definite LRTI cases were viral, reflecting the typical microbiology of pediatric community-acquired LRTI.(5) Etiologies of non-infectious respiratory failure included seizures, trauma, airway abnormalities, and ingestion. Differential gene expression analysis between Definite and No Evidence subjects identified 4,718 genes at a Benjamini-Hochberg adjusted p-value (P_adj_)<0.05, with the gene fatty acid-binding protein 4 (*FABP4*) emerging as the most promising candidate biomarker (P_adj_<0.0001) (Figure 1A). Five-fold cross-validation on normalized *FABP4* gene expression in the Definite and No Evidence groups yielded an area under the receiver operator curve (AUC) of 0.90±0.07 for diagnosing LRTI, equating to a sensitivity of 86±7% and specificity of 90±7% at Youden’s index (Figure 1B). *FABP4* classified 73.7% of the Suspected group and 54.1% of the Indeterminate group as LRTI, aligning with stronger suspicion of LRTI in the former.

**Table 1:**
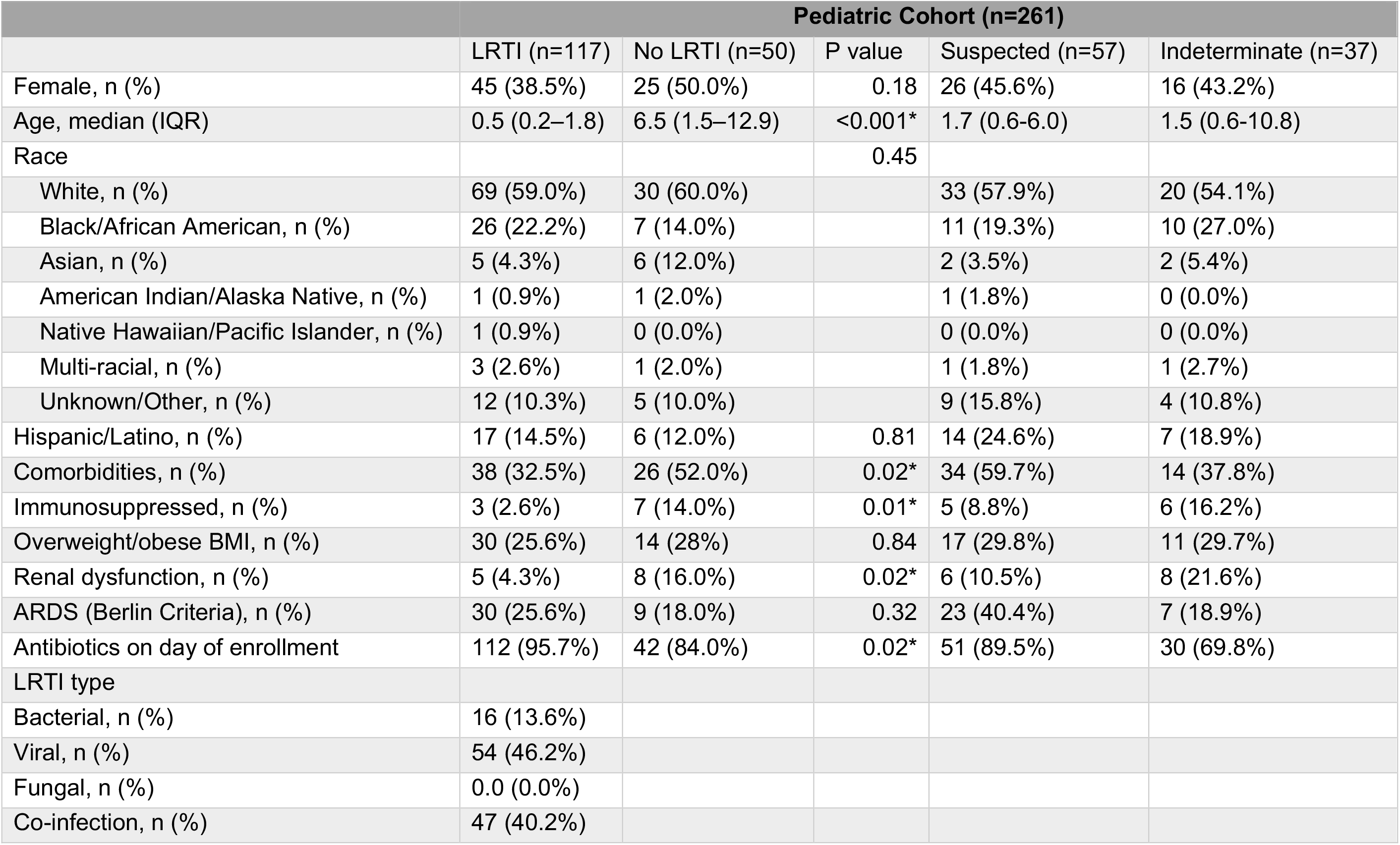
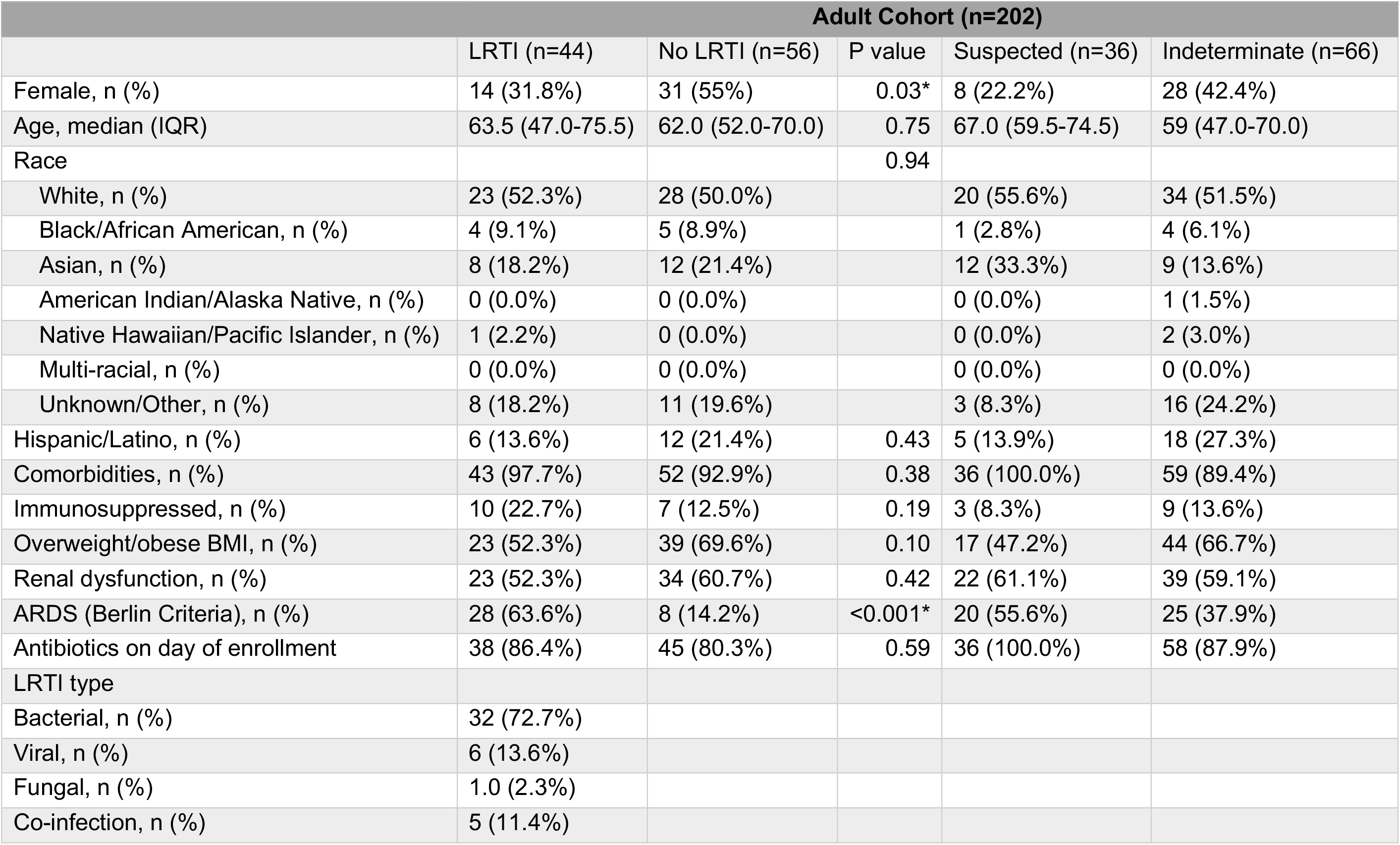
Demographic and clinical characteristics of the pediatric and adult cohorts. Abnormal BMI was classified as >25 kg/m^2^ for adults and >85^th^ percentile for children. Renal dysfunction was classified as creatinine above the upper limit of normal and/or need for renal replacement therapy while hospitalized. Mann-Whitney test was used for all continuous variables, and Fisher’s exact test was used for all categorical values. Asterisk (*) indicates p<0.05 between the Definite and No Evidence groups. ARDS, acute respiratory distress syndrome.

| Pediatric Cohort (n=261) |  |  |  |  |  |
| --- | --- | --- | --- | --- | --- |
|  | LRTI (n=117) | No LRTI (n=50) | P value | Suspected (n=57) | Indeterminate (n=37) |
| Female, n (%) | 45 (38.5%) | 25 (50.0%) | 0.18 | 26 (45.6%) | 16 (43.2%) |
| Age, median (IQR) | 0.5 (0.2–1.8) | 6.5 (1.5–12.9) | <0.001* | 1.7 (0.6-6.0) | 1.5 (0.6-10.8) |
| Race |  |  | 0.45 |  |  |
| White, n (%) | 69 (59.0%) | 30 (60.0%) |  | 33 (57.9%) | 20 (54.1%) |
| Black/African American, n (%) | 26 (22.2%) | 7 (14.0%) |  | 11 (19.3%) | 10 (27.0%) |
| Asian, n (%) | 5 (4.3%) | 6 (12.0%) |  | 2 (3.5%) | 2 (5.4%) |
| American Indian/Alaska Native, n (%) | 1 (0.9%) | 1 (2.0%) |  | 1 (1.8%) | 0 (0.0%) |
| Native Hawaiian/Pacific Islander, n (%) | 1 (0.9%) | 0 (0.0%) |  | 0 (0.0%) | 0 (0.0%) |
| Multi-racial, n (%) | 3 (2.6%) | 1 (2.0%) |  | 1 (1.8%) | 1 (2.7%) |
| Unknown/Other, n (%) | 12 (10.3%) | 5 (10.0%) |  | 9 (15.8%) | 4 (10.8%) |
| Hispanic/Latino, n (%) | 17 (14.5%) | 6 (12.0%) | 0.81 | 14 (24.6%) | 7 (18.9%) |
| Comorbidities, n (%) | 38 (32.5%) | 26 (52.0%) | 0.02* | 34 (59.7%) | 14 (37.8%) |
| Immunosuppressed, n (%) | 3 (2.6%) | 7 (14.0%) | 0.01* | 5 (8.8%) | 6 (16.2%) |
| Overweight/obese BMI, n (%) | 30 (25.6%) | 14 (28%) | 0.84 | 17 (29.8%) | 11 (29.7%) |
| Renal dysfunction, n (%) | 5 (4.3%) | 8 (16.0%) | 0.02* | 6 (10.5%) | 8 (21.6%) |
| ARDS (Berlin Criteria), n (%) | 30 (25.6%) | 9 (18.0%) | 0.32 | 23 (40.4%) | 7 (18.9%) |
| Antibiotics on day of enrollment | 112 (95.7%) | 42 (84.0%) | 0.02* | 51 (89.5%) | 30 (69.8%) |
| LRTI type |  |  |  |  |  |
| Bacterial, n (%) | 16 (13.6%) |  |  |  |  |
| Viral, n (%) | 54 (46.2%) |  |  |  |  |
| Fungal, n (%) | 0.0 (0.0%) |  |  |  |  |
| Co-infection, n (%) | 47 (40.2%) |  |  |  |  |

| Adult Cohort (n=202) |  |  |  |  |  |
| --- | --- | --- | --- | --- | --- |
|  | LRTI (n=44) | No LRTI (n=56) | P value | Suspected (n=36) | Indeterminate (n=66) |
| Female, n (%) | 14 (31.8%) | 31 (55%) | 0.03* | 8 (22.2%) | 28 (42.4%) |
| Age, median (IQR) | 63.5 (47.0-75.5) | 62.0 (52.0-70.0) | 0.75 | 67.0 (59.5-74.5) | 59 (47.0-70.0) |
| Race |  |  | 0.94 |  |  |
| White, n (%) | 23 (52.3%) | 28 (50.0%) |  | 20 (55.6%) | 34 (51.5%) |
| Black/African American, n (%) | 4 (9.1%) | 5 (8.9%) |  | 1 (2.8%) | 4 (6.1%) |
| Asian, n (%) | 8 (18.2%) | 12 (21.4%) |  | 12 (33.3%) | 9 (13.6%) |
| American Indian/Alaska Native, n (%) | 0 (0.0%) | 0 (0.0%) |  | 0 (0.0%) | 1 (1.5%) |
| Native Hawaiian/Pacific Islander, n (%) | 1 (2.2%) | 0 (0.0%) |  | 0 (0.0%) | 2 (3.0%) |
| Multi-racial, n (%) | 0 (0.0%) | 0 (0.0%) |  | 0 (0.0%) | 0 (0.0%) |
| Unknown/Other, n (%) | 8 (18.2%) | 11 (19.6%) |  | 3 (8.3%) | 16 (24.2%) |
| Hispanic/Latino, n (%) | 6 (13.6%) | 12 (21.4%) | 0.43 | 5 (13.9%) | 18 (27.3%) |
| Comorbidities, n (%) | 43 (97.7%) | 52 (92.9%) | 0.38 | 36 (100.0%) | 59 (89.4%) |
| Immunosuppressed, n (%) | 10 (22.7%) | 7 (12.5%) | 0.19 | 3 (8.3%) | 9 (13.6%) |
| Overweight/obese BMI, n (%) | 23 (52.3%) | 39 (69.6%) | 0.10 | 17 (47.2%) | 44 (66.7%) |
| Renal dysfunction, n (%) | 23 (52.3%) | 34 (60.7%) | 0.42 | 22 (61.1%) | 39 (59.1%) |
| ARDS (Berlin Criteria), n (%) | 28 (63.6%) | 8 (14.2%) | <0.001* | 20 (55.6%) | 25 (37.9%) |
| Antibiotics on day of enrollment | 38 (86.4%) | 45 (80.3%) | 0.59 | 36 (100.0%) | 58 (87.9%) |
| LRTI type |  |  |  |  |  |
| Bacterial, n (%) | 32 (72.7%) |  |  |  |  |
| Viral, n (%) | 6 (13.6%) |  |  |  |  |
| Fungal, n (%) | 1.0 (2.3%) |  |  |  |  |
| Co-infection, n (%) | 5 (11.4%) |  |  |  |  |

**Figure 1:**
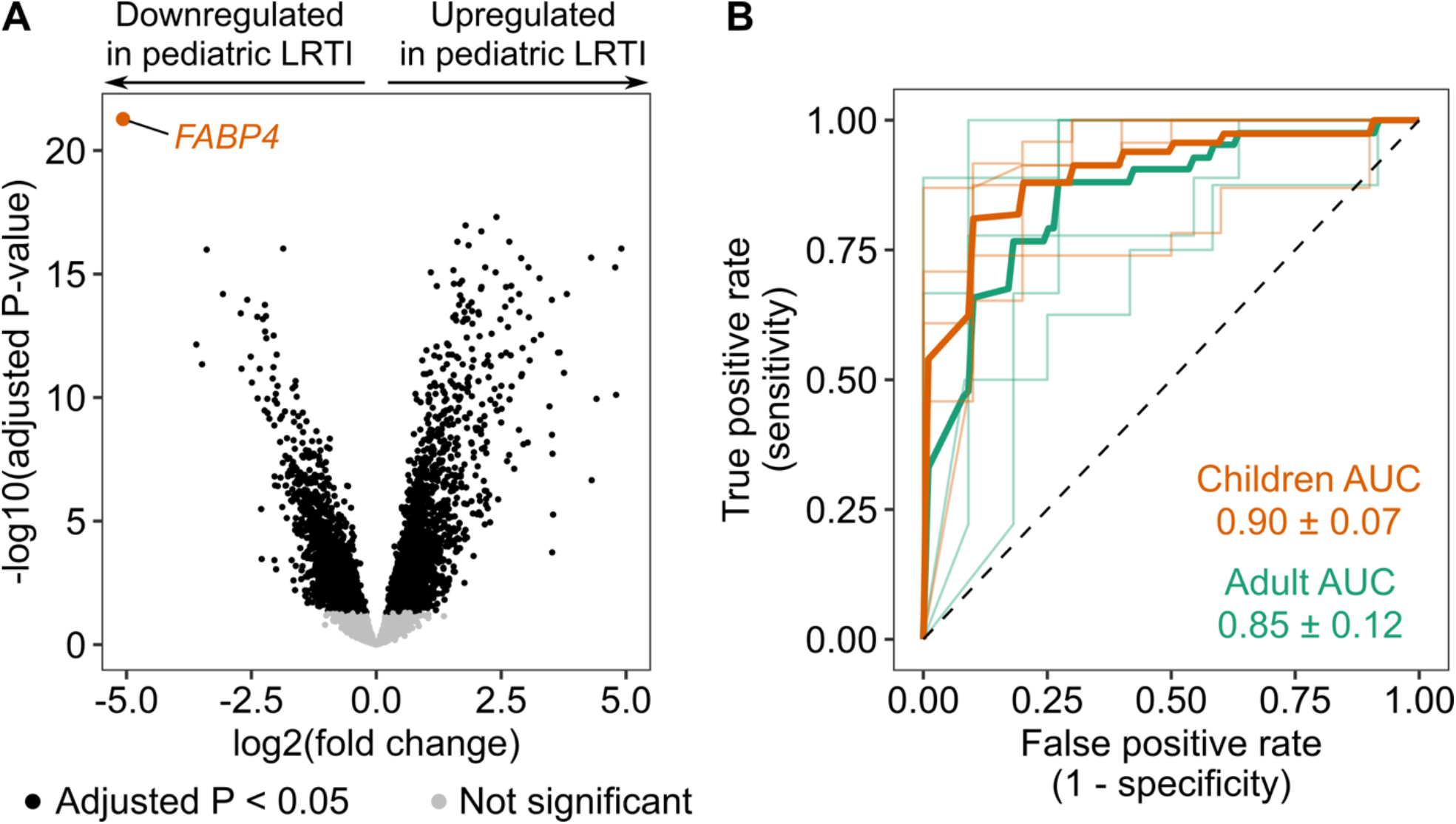
*FABP4* performance in adult and pediatric cohorts. (A) Volcano plot displaying differential expression of all genes quantified by RNA sequencing between Definite LRTI and No Evidence subjects within the pediatric cohort. Log2(fold change) is plotted against -log10(P_adj_) for each gene. Statistically significant genes (P_adj_<0.05) are shown in black and non-significant genes are shown in grey. *FABP4* gene is highlighted in orange as highly downregulated in LRTI patients. (B) Receiver operating characteristic (ROC) curve demonstrating *FABP4* performance in diagnosing LRTI in the pediatric (orange) and adult (green) cohorts. The faded lines indicate results from each of the 5-fold validations on normalized *FABP4* gene counts. The area under the curve (AUC) is presented as mean ± standard deviation.

Given that LRTI microbiology and immune responses vary with age, it was uncertain whether *FABP4* would perform as well in diagnosing adult LRTI. We therefore assessed *FABP4* expression in an independent prospective cohort of 202 adults requiring intubation for acute respiratory failure (Table 1).(3) A similar retrospective clinical LRTI adjudication approach identified 44 Definite, 36 Suspected, 66 Indeterminate, and 56 No Evidence subjects. In contrast to the pediatric cohort, the adult Definite subjects had predominantly bacterial LRTI and included subjects with both hospital-acquired and community-acquired pneumonia. Etiologies of non-infectious respiratory failure included cardiac disease, neurologic syndromes, and primary pulmonary processes (e.g., COPD, ILD, pulmonary embolism). Utilizing transcriptional profiling of TA samples collected <72 hours after intubation, we compared *FABP4* gene expression across LRTI status. *FABP4* exhibited a similarly striking decrease in expression in adults with Definite LRTI compared to No Evidence (P_adj_<0.0001) and achieved an AUC of 0.85±0.12 when used as a diagnostic classifier in these two groups (Figure 1). Similar to the pediatric cohort, *FABP4* predicted LRTI in 66.7% of the Suspected group and 31.8% of the Indeterminate group.

So what is *FABP4*, and why might it be downregulated in LRTI? Fatty-acid binding proteins are a family of intracellular proteins that regulate fatty acid trafficking and signaling.(6) In the lung, *FABP4* is considered a marker of tissue-resident alveolar macrophages, and severe SARS-CoV-2 infection has recently been shown to drastically deplete the *FABP4-*expressing alveolar macrophage population.(7,8) Prior studies have also linked increased *FABP4* levels to non-infectious chronic pulmonary conditions including bronchopulmonary dysplasia, sarcoidosis, and asthma.(6) The precise mechanism for *FABP4* downregulation in infection – or, alternatively, upregulation in non-infectious lung disease – remains unclear. From a diagnostic lens, a study of ventilator-associated pneumonia identified FABP4 as one of the most downregulated proteins in TA, and a study of COPD patients noted decreased sputum FABP4 protein levels in bacterial infection, supporting our transcriptomic findings at the protein level.(9,10)

Taken together, these results demonstrate that *FABP4* is a promising age-agnostic, inverse-biomarker for all-cause LRTI in two independent cohorts of critically ill, intubated patients. *FABP4* achieved high accuracy for LRTI diagnosis in both cohorts, with performance exceeding those of existing clinical biomarkers(1,2) despite differing ages, microbiology, comorbidity burden, and preceding healthcare exposures. As host immune responses are more dynamic and informative at the site of infection, the identification of an accurate LRTI biomarker from respiratory fluid rather than peripheral blood, while novel, is perhaps not surprising. Diagnostically, *FABP4* could add value to standard TA culture, both in discriminating LRTI pathogens from colonizing microbes and in diagnosing LRTI when cultures are hindered by preceding antibiotics. In addition to improving LRTI diagnosis for the individual patient, a pan-pathogen host biomarker of LRTI could have utility for pandemic preparedness and infection control, enabling rapid screening, early isolation, and other prevention measures.

These preliminary findings require independent validation in additional cohorts with both transcriptomic and proteomic approaches. It remains unknown whether *FABP4* has diagnostic utility in non-critically ill patients, or when measured in more broadly accessible sample types, such as sputum, nasal swabs, or blood. To better understand the mechanism of *FABP4* modulation and to gauge whether particular cell types may mediate or confound *FABP4* expression, cytometry and single-cell RNA sequencing on lower respiratory tract samples from patients with LRTI, non-infectious respiratory syndromes, and healthy controls will be valuable. Lastly, an important step towards clinical translation will be development of RT-qPCR and/or lateral-flow protein assays. In sum, we find that pulmonary *FABP4* is a novel, downregulated biomarker of severe LRTI across the age spectrum, with promising utility in critically ill patients.

## Data Availability

All data, code, and results are available at: https://github.com/infectiousdisease-langelier-lab/LRTI_FABP4_classifier for reviewing and reproducing the analyses.

## Ethics Statement

The pediatric cohort study was approved by the Collaborative Pediatric Critical Care Research IRB at the University of Utah (protocol #00088656). Informed consent was obtained from parents or other legal guardians. The adult cohort was approved by the University of California Institutional Review Board (protocol #10-02701) and informed consent was obtained from patients or surrogate decision makers.

## Notes

### Competing Interest Statement

The authors have declared no competing interest.

